# Epistatic SNP network analysis (ESNA): A scalable framework for genome-wide detection of higher-order genetic interactions

**DOI:** 10.64898/2026.05.08.26352667

**Authors:** Yang Zhang, Ming Han, Amirthagowri Ambalavanan, Danai Topouza, Zhi Yi Fang, Sara A. Stickley, Shrey Anand, Stuart Turvey, Piushkumar J. Mandhane, Elinor Simons, Theo J. Moraes, Padmaja Subbarao, Jihoon Choi, Qingling Duan

## Abstract

Although genome-wide association studies (GWASs) have been widely applied to investigate the genetic basis of common traits and diseases in human populations, the associated loci do not fully account for the estimated heritability. The missing heritability may be explained, in part, by epistasis or gene-gene interactions. Existing methods for detecting epistasis, however, are limited to pair-wise interactions and/or targeted genomic regions. Here, we present a novel model, termed the *Epistatic SNP Network Analysis* (*ESNA*), which detects higher-order epistatic interactions using genome-wide SNP data. ESNA employs a scale-free network algorithm within a parallel computing framework that identifies modules of correlated SNPs, potentially interacting variants that converge on common biological pathways, while enhancing computational efficiency. We applied *ESNA* to investigate epistatic interactions contributing to respiratory outcomes such as recurrent wheeze and asthma among preschool-aged children in the CHILD Cohort Study. Using genome-wide data comprising 775,569 SNPs from 1,899 children, *ESNA* identified 914 SNP network modules, 9 of which were significantly associated with recurrent wheeze between ages 2 and 5 years (P<5.47×10^-5^). Furthermore, 7 of these wheeze-associated modules were also associated with asthma by age 5 years (P<5.47×10^-5^). Pathway enrichment analysis revealed that the associated modules consist of SNPs located in genes previously implicated in asthma and related biological processes, such as cellular response to stimuli and nervous system development. Compared to existing network-based methods for epistasis, *ESNA* demonstrated substantial improvements in computational efficiency, reducing memory usage by 50% and processing genome-wide SNP data 48 times faster. The code implementation and documentation are available at https://github.com/ComputationalGenomicsLaboratory/ESNA.

## Introduction

Genome-wide association studies (GWASs) identify genetic variants linked to common traits and complex diseases [1] by testing hundreds of thousands to millions of single nucleotide polymorphisms (SNPs) for association with phenotypes. While GWASs have revealed loci for numerous diseases such as asthma, obesity, inflammatory bowel disease, and schizophrenia, these variants explain only a small fraction of the heritability, which is the proportion of trait variation due to genetic differences [2]. The “missing heritability” may be attributed, in part, to gene–gene interactions known as epistasis [3].

Epistasis describes non-additive interactions where the effect of one locus depends on another [4, 5]. Detection methods are either hypothesis-driven, focused on biologically plausible candidates, or hypothesis-free, scanning genome-wide for novel interactions [3]. Detecting higher-order epistasis is computationally challenging, with complexity increasing exponentially by interaction order [3, 4, 6]. For example, 2 million SNPs yield 2x10^12^ pairwise and 1.3x10^18^ three-way interactions, with n-way interaction scaling as (2^n^x10^6n^)/(n!) [7]. A full genome-wide search for three-way interactions requires hundreds of petabytes of memory and storage, which may be prohibitive for many research environments.

Current methods such as *GPU-Based Linear Regression for Detection of Epistasis* (*GLIDE*) detect only pairwise interactions [8-13], missing more complex patterns. The *Weighted Interaction SNP Hub* (*WISH*) package applies a scale-free network algorithm using multi-core processing but is designed for targeted regions instead of genome-wide data [13]. Recent approaches capture higher-order interactions genome-wide such as *epiMEIF*, which employs mixed-effect forests to identify multi-locus dependencies [14]; a transformer-based deep learning model by *Graça et al.* (2024) [15]; and *Epi-SSA* [16], which leverages evolutionary search algorithms. While innovative, these methods require large, labeled datasets and substantial computational overhead, which constrain the ability to jointly model local and global interaction structures.

Network-based approaches offer a complementary perspective. For example, Module Detection via SNP Networks (*MDSN*) constructs SNP-SNP interaction networks from fast pairwise association scores and identify phenotype-associated clusters [17]. However, the reliance on pairwise scores limits the detection of complex higher-order patterns. Additionally, its CPU-based implementation lacks memory and has been tested only on small datasets, constraining its scalability to genome-wide applications.

To address these limitations of existing methods, we developed Epistatic SNP Network Analysis (*ESNA*), a network-based model for large-scale epistasis detection. *ESNA* uses a parallelized computing framework to distribute tasks across multiple high-performance computing (HPC) clusters and cores. Partitioning genome-wide SNP data and applying a customized parallel algorithm enables the efficient detection of higher-order SNP interactions. Building on prior tools such as *GLIDE* and *WISH*, *ESNA* is, to our knowledge, the first network-based framework for detecting higher-order epistatic interactions at a genome-wide scale. We applied *ESNA* to genome-wide SNP data from the Canadian CHILD Cohort Study [18], which uncovered genetic interactions that may contribute to childhood respiratory outcomes.

## Materials and Methods

### CHILD Cohort and Genomics Data

To evaluate *ESNA* for detecting interaction effects of SNPs and its computational efficiency, we applied our novel framework to genome-wide SNP data from the CHILD Cohort Study. This large, prospective birth cohort recruited 3,542 pregnant women and their offspring to investigate early-life determinants of chronic childhood diseases, including asthma and related outcomes [18]. Health questionnaires were administered biannually between ages 2 and 3 years and annually thereafter until age 5, capturing parent-reported wheeze frequency and triggers [19]. These responses were used to define recurrent wheeze as two or more wheezing episodes in a single year between ages 2 and 5 years. Wheezing episodes prior to age 2 were excluded due to their higher likelihood of being transient or infection-related [20]. Recurrent wheeze during early childhood was selected as the primary respiratory outcome, as it is a key predictor of asthma and long-term pulmonary impairment [21]. Asthma was assessed at age 5 years by a specialist and included as a secondary phenotype given that diagnosis in preschool-aged children is challenging [22, 23], due to disease heterogeneity [24, 25].

### Genomic Data Preprocessing

Genomic DNA was extracted from cord blood and genotyped using the Illumina HumanCoreExome BeadChip, yielding 557,006 SNPs across 2,967 children [26, 27]. Standard quality control (QC) procedures were conducted in *PLINK* [28] following previously published pipelines [29, 30], which excluded a total of 132 individuals for any of the following criteria: sex discrepancies, high heterozygosity, genotype missingness, or relatedness. Genotype imputation was performed via the Michigan Imputation Server using the Haplotype Reference Consortium (HRC r1.1 2016), resulting in approximately 28 million variants per individual. We retained common variants with a minor allele frequency (MAF) ≥ 0.05, yielding 5,475,011 SNPs for downstream analysis.

To reduce redundancy, we applied linkage disequilibrium (LD) pruning (*r*^2^ > 0.6), producing a final dataset of 775,613 SNPs across 1,899 children who also had phenotype data. Of these, 1,005 were male and 894 were female. A total of 268 children had recurrent wheeze between ages 2-5 years, while 1,631 others served as non-wheezing controls. Fewer children (n=145) were diagnosed with asthma by age 5, with 1,754 classified as non-asthmatic for analysis.

### ESNA Model Overview

The overall ESNA workflow is illustrated in **Figure 1A**, comprising four major steps: 1) pairwise SNP-SNP epistatic analysis using GLIDE [12] to generate an interaction-based similarity matrix; 2) construction of a global SNP network (GSN) by transforming the similarity matrix into a denoised, higher-order interaction network; 3) unsupervised clustering of SNPs into network modules based on GSN topology; and 4) phenotype association testing of modules using the Sequence Kernel Association Test (SKAT) [31], followed by biological annotation of significant modules.

**Figure 1.**
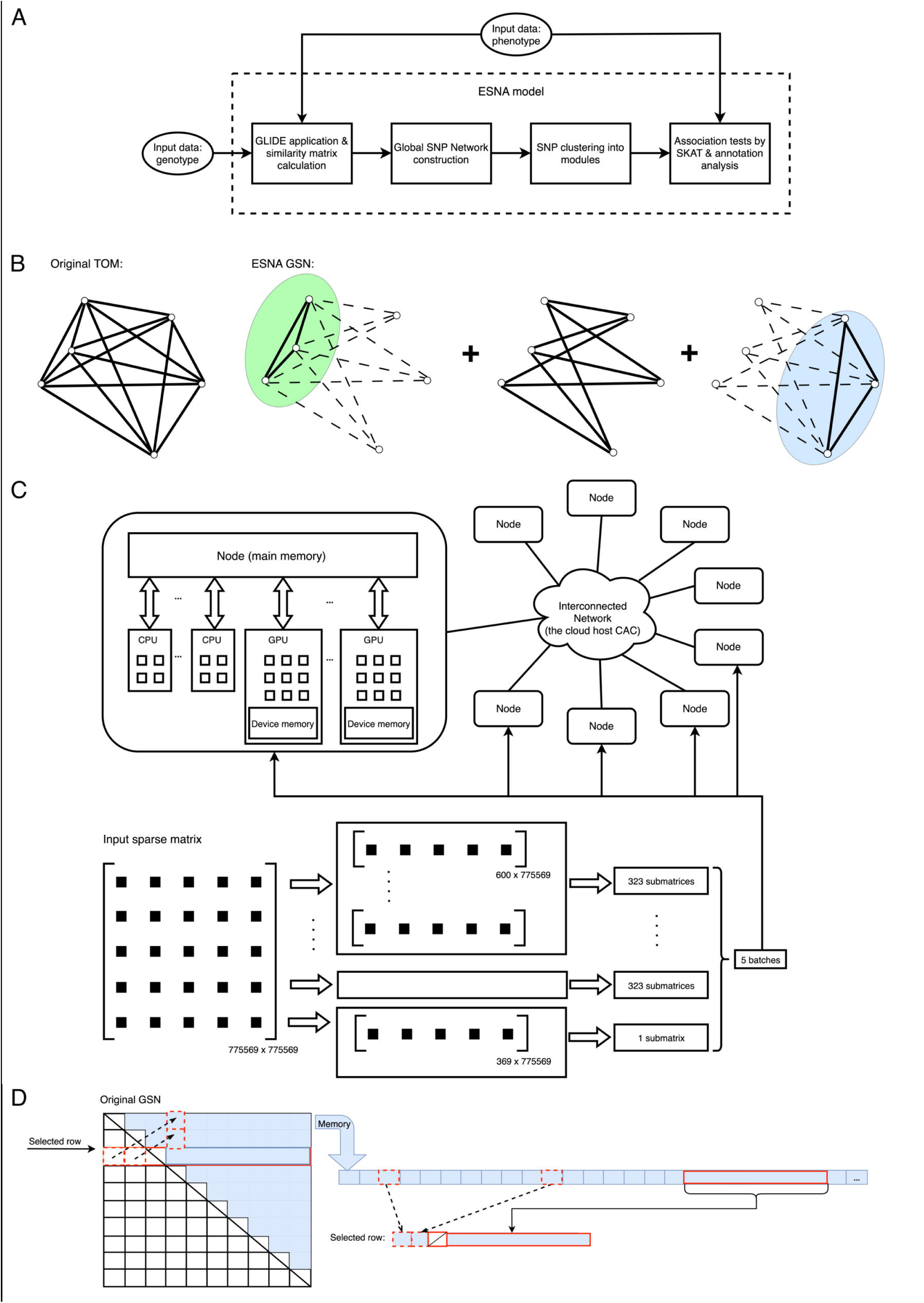
Overview of ESNA and construction of the Global SNP Network (GSN). **(A)** The complete ESNA workflow for genome-wide epistasis analysis. Input data include genotypes (e.g., SNP arrays) and phenotypes (e.g., recurrent wheeze, asthma). ESNA proceeds through four computational steps: (1) pairwise SNP-SNP epistatic interactions are computed using GLIDE to produce a similarity matrix; (2) similarity scores are denoised and transformed into a higher-order Global SNP Network (GSN); (3) SNPs are clustered into modules based on topological overlap in the GSN; and (4) the resulting modules are tested for phenotype association using SKAT, followed by biological annotation. **(B)** Schematic of GSN construction via block-wise topology. ESNA partitions the genome-wide SNP similarity matrix into subnetworks (blocks), where both intra-block connectivity (shown in green and blue) and inter-block neighbor contributions (dashed lines) are integrated to define higher-order relationships. This design enables scalable and structure-aware modeling of epistatic architecture. **(C)** *GPU-accelerated parallel block-wise computation for* large-scale *GSN Matrix.* The adjacency matrix (775,569 × 775,569) was partitioned into five blocks, each containing 323 submatrices with dimensions 600 × 775,569, along with one smaller tail block (369 × 775,569), forming five batches that were distributed across five GPU-enabled compute nodes. Each node processed its assigned submatrices in parallel, enabling efficient network construction without exceeding memory limits. **(D)** QuickMap strategy for memory-efficient matrix storage. The full upper triangle of the GSN matrix was compressed into a 1-Dimension (1D) array, enabling on-demand reconstruction of any SNP row. For a selected row (red box), its corresponding interaction values are derived from (1) preceding elements before the diagonal, (2) the diagonal entry, and (3) elements beyond the diagonal (blue bracketed area).

Steps 1 and 4 leverage established tools (*GLIDE* and *SKAT*), whereas steps 2 and 3 represent the novel methodological contributions of *ESNA*. In particular, *ESNA* employs a computationally efficient framework for converting pairwise SNP interactions into a topological structured network and detecting epistatic modules of potential biological relevance. These innovations address core computational limitations of previous approaches including memory usage, data scale, and algorithmic extensibility that have hindered genome-wide epistasis analysis.

### Pairwise SNP-SNP Epistatic Analysis

To identify epistatic interactions among genetic variants, we employed the *GLIDE* package *[12]* to compute pairwise interaction effects among all SNPs. *GLIDE* applies GPU-accelerated linear regression to test each SNP pair for interaction effects on the phenotypes (e.g., recurrent wheeze), producing a t-statistic for the interaction term of every SNP pair. We applied a t-score threshold of > 3.0 (as recommended by *GLIDE*) to filter out negligible interactions and reduce input data size, thereby retaining only the strongest epistatic signals. The surviving pairwise interaction scores were converted into a symmetric SNP-SNP similarity matrix of dimension *N* × *N* (where *N* is the number of SNPs). Each entry of this matrix represents the strength of epistatic association between two SNPs, with higher t-scores indicating a stronger potential interaction effect on the phenotype. This step yielded a sparse matrix of pairwise connections that served as the input for subsequent network analysis.

### Global SNP Network (GSN) Construction

To capture higher-order epistatic structure beyond isolated pairwise interactions, we transformed the SNP similarity matrix into a global SNP network (GSN). First, we converted the pairwise t-scores into a weighted adjacency matrix through an exponential soft-thresholding method. Specifically, each pairwise similarity value was rescaled in a logarithmic domain [32] and raised to a power *λ* (a tunable parameter in Equation 1) to accentuate strong interactions. The power *λ* was chosen according to the scale-free topology criterion of network construction [35], ensuring that the distribution of SNP connectivity in the resulting network approximated a scale-free network. A normalization constant *β* was applied so that the adjacency weights remained in a (0,1) range. This yielded an adjacency matrix *A* = [*a*_*ij*_], where a higher *a*_*ij*_ indicates a stronger direct epistatic connection between SNP_i_ and SNP_j_.

Next, we constructed the GSN by calculating a topological overlap measure for each SNP pair, analogous to the Topological Overlap Matrix (TOM) used in weighted gene co-expression networks (*WGCNA*) framework [35]. This step incorporates not only the direct adjacency between two SNPs but also the extent to which they share interaction partners (neighbors) in common. For each pair of SNPs *i* and *j*, the GSN weight *w*_*ij*_ in subnetwork block *GSN*_*i*∈*N*_*k*_,*j*_ was defined as follows:

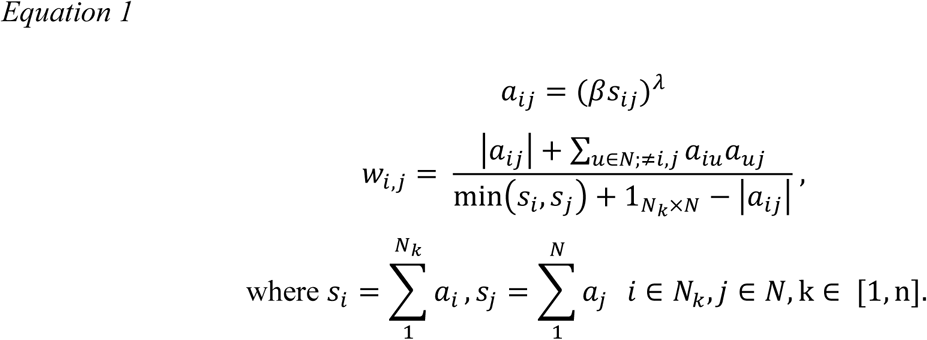

In Equation 1, *a*_*ij*_ denotes adjacency between SNP_i_ and SNP_j_, derived from the similarity score *s*_*ij*_ via an exponential denoising transformation. The magnitude |*a*_*ij*_| reflects the strength of this adjacency. The term *w*_*i*,*j*_ represents elements in the *k*-th subnetwork block of the *GSN*, where [*w*_*i*,*j*_] = *GSN*_*i*,*j*_ with dimensions *N*_*k*_ × *N*. Here, k ∈ [1, n], while *n* indicates the maximum number of network blocks.

The numerator of Equation 1, ∑_*u*∈*N*_*k*_;≠*i*,*j*_ *a*_*iu*_*a*_*uj*_, quantifies the cumulative strength and number of shared neighbors between SNPs *i* and *j*, capturing indirect epistatic interactions via common connectivity. The denominator aggregates the contributions of all neighbors for both SNPs *i* and *j*, serving as a normalization factor that scales the overlap score to the [0, 1]. Terms 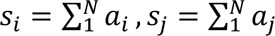 represent the overall connectivity of SNP_i_ and SNP_j_, respectively.

By construction, *w*_*i*,*j*_ ≈ 1 when *i* = *j* or when two SNPs share all the same neighbors (complete topological overlap), whereas *w*_*i*,*j*_ ≈ 0 indicates little to no shared interactions. This formulation captures both direct and indirect relationships, enabling the network to reflect higher-order epistatic structure.

Constructing the GSN naïvely over the full *N* × *N* matrix is computationally infeasible for genome-wide datasets. To address this, we implemented a block-wise parallelization strategy. The adjacency matrix was divided into disjoint row-wise subnetwork blocks, and Equation 1 was computed in parallel across these blocks, with block size determined by available computational resources. This approach avoids the need to store or multiply the full matrix in memory, allowing scalable application to large SNP datasets.

To account for interactions spanning across blocks, we incorporated inter-block information by averaging contributions from neighboring SNPs located in other blocks. This ensured that each subnetwork block captured both local and global connectivity, yielding results equivalent to a single global TOM calculation but with significantly reduced memory and runtime demands.

After assembling all subnetwork outputs, the final GSN was constructed as the union of all blocks:

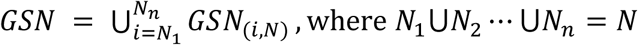

Notably, the denoising and neighbor-incorporation steps resulted in a fully dense matrix, where every SNP pair had a non-zero topological overlap score. This density reflected the pervasive connectivity of epistatic relationships and introduced significant computational challenges for downstream analysis. To address this, we implemented a tailored clustering strategy to efficiently identify SNP modules from the GSN, as described in the next step.

### Clustering of SNP Modules

To identify modules of highly interconnected SNPs from the GSN, we employed a two-stage clustering strategy combining parallel K-means pre-clustering with hierarchical refinement using the *DynamicTreeCut* algorithm [33-35].

In the first stage, we applied parallelized K-means clustering to partition SNPs into preliminary groups, based on their pairwise similarity within the GSN. We initialized centroids randomly and iteratively reassigned SNPs to the most similar centroid (highest similarity e.g., Euclidean Distance). After each iteration, centroids were updated by averaging the network profiles of all SNPs within a cluster, and the reassignment step repeated. This process parallelized the computation across SNPs in the GSN and terminated upon convergence or reaching a predefined iteration limit. The resulting preliminary clusters captured dense local connectivity within the GSN.

In the second stage, we refined these clusters using hierarchical clustering followed by the *DynamicTreeCut* algorithm [35]. A dendrogram was constructed for each preliminary cluster using a distance metric derived from topological overlap. *DynamicTreeCut* adaptively determined the final module boundaries by analyzing the dendrogram structure, avoiding the need for a fixed height threshold. This approach merged or split branches derived from K-means cluster members based on data-driven criteria, improving cluster robustness and granularity.

By integrating fast, parallel K-means pre-clustering with adaptive hierarchical refinement, we generated a final set of SNP modules, each comprising SNPs with high topological overlap. These modules served as the input for downstream phenotype association testing.

### Sequence Kernel Association Tests and Functional Annotation

Following module identification, we assessed each SNP module’s association with the phenotype of interest using a set-based statistical framework. Specifically, we applied the *Sequence Kernel Association Test* (*SKAT*) [31], a variance-component method originally developed for rare variant association studies, to evaluate the collective effect of all SNPs within each module. This approach accommodates correlation among SNPs within a module and tests whether their aggregate additive effect is significantly associated with the phenotype.

To improve statistical power, we incorporated per-SNP weighting into the *SKAT* model based on external evidence of SNP relevance. We derived weights from the absolute effect sizes (beta coefficients) reported in a large, independent asthma GWAS by *Pividori et al.* [36]. SNPs with larger effects in the external study were assigned higher weights, reflecting a prior expectation of stronger phenotypic impact. This weighting strategy enhanced sensitivity to detect associations within modules that harbor known risk loci, while still accommodating discovery of novel epistatic effects.

We conducted a conventional single-SNP genome-wide association study (GWAS) using *PLINK* [28], applying logistic regression under an additive model and filtering for common variants (minor allele frequency > 5%). This baseline analysis enabled comparison between *ESNA*’s module-level results and traditional single-marker associations, allowing us to evaluate *ESNA*’s capacity to uncover polygenic interaction signals potentially overlooked by standard GWAS.

To interpret the biological relevance of associated SNP modules, we performed functional annotation and pathway enrichment analysis. All SNPs within each module were annotated using the *Ensembl Variant Effect Predictor (VEP)* tool for the reference genome build GRCh37 [37], providing information on genomic context (e.g., intronic, intergenic, coding) and predicted functional consequences (e.g., missense, regulatory, splice-altering). For enrichment analysis, we used *g-Profiler* [38] to test whether SNPs mapped to genes overrepresented in known biological pathways, including Gene Ontology categories and disease-relevant processes.

Together, the integration of SKAT-based association testing, variant annotation, and pathway enrichment provided both statistical and biological insights, enabling prioritization of SNP modules with plausible mechanistic roles in complex traits such as asthma and recurrent wheezing.

### Computational Implementation

Figure 1 summarizes how ESNA leverages both network topology and distributed computation to enable efficient analysis of genome-wide SNP interaction data. Figure 1B-D illustrates key aspects of the computational framework underpinning *ESNA*. Figure 1B provides a conceptual visualization of the Global SNP Network (GSN), highlighting how topological overlap relationships among SNPs are constructed from pairwise similarity values to capture higher-order epistatic interactions. Figure 1C presents a computational schematic representation of the GSN construction algorithm in parallel block processing, implements the procedure defined in **Equation 1**. We implemented a custom indexing utility, QuickMap, which stores only the upper triangular portion of the GSN matrix in a one-dimensional array and reconstructs any row on demand. This approach reduces memory usage by approximately 50% for non-sparse GSN data and represents a purpose-built solution for large-scale SNP network storage within the ESNA framework (Figure 1D).

### Software and Computing Environment

The *ESNA* methodology was implemented in Python 3.8 (with key scientific libraries) and executed on a HPC cluster running Linux (CentOS 7). All analyses were conducted on the Centre for Advanced Computing cloud cluster at Queen’s University, Canada [39], which provided multi-node CPU and GPU capabilities (CUDA 11.1). *GLIDE* (the pairwise epistasis analysis in step 1) was run on NVIDIA Tesla V100 GPUs, enabling rapid calculation on the order of *N*^2^pairwise SNP interactions in parallel. Subsequent SNP network construction and clustering computations were distributed across multiple CPU cores and cluster nodes as detailed below.

### Data Structures and Memory Optimization

Genome-wide epistasis analysis generates extremely large matrices, so we employed several strategies to efficiently handle data storage and processing. We used the Hierarchical Data Format (HDF5 [40]) to store intermediate results (such as the sparse SNP similarity matrix from *GLIDE*) on disk storage. HDF5 is optimized for large datasets, allowing structured storage and fast I/O access, and thus was well-suited to manage the tens of billions of pairwise interaction values. In memory, we took advantage of sparse matrix representations from SciPy (*scipy.sparse*) to store the *GLIDE* output matrix, since only a fraction of SNP pairs surpassed the t-score threshold. Using sparse matrix formats such as coordinate (COO) and list-of-lists (LIL) [41], which kept only non-zero interaction entries in memory, significantly reducing RAM usage [42]. We further minimized memory overhead by using 32-bit floating point precision (float32) for storing similarity and network values, instead of 64-bit doubles. This halved the memory required for matrix storage while retaining sufficient numeric precision for our calculations. In aggregate, these choices (HDF5 storage, sparse matrices, and float32 precision) allowed us to handle the initial pairwise similarity matrix (which would contain, if fully dense, on the order of 6 ∗ 10^11^ entries for around 775k SNPs) in a feasible manner on the HPC infrastructure.

### Parallel and GPU-accelerated Computing

We integrated both multi-core CPU parallelism and GPU acceleration to speed up the computationally intensive steps of ESNA. Construction of the GSN (Equation 1) and SNP clustering were parallelized across SNP blocks and distributed over multiple compute nodes. We implemented the parallelism using the Message Passing Interface (MPI) via the *mpi4py* Python library [33], enabling SNP blocks to be processed both across multiple nodes (inter-node) and across CPU cores within a single node (intra-node) concurrently. Within each node, computations were further accelerated using GPUs and optimized libraries. We leveraged *CuPy* [43], a GPU-accelerated array library compatible with NumPy, to perform large matrix operations (such as sparse matrix multiplication and element-wise transformations) on NVIDIA GPUs. For custom operations not directly supported by *CuPy*, we used *numba.cuda* just-in-time (JIT) compiler [44] to offload Python code to the GPU, achieving low-level performance for those routines. This combination of GPU computing and parallel CPU processing (a form of hybrid computing) was key to constructing the GSN and SNP clustering, which required demanding matrix computations. For example, instead of computing a massive matrix multiplication for the entire TOM in one step (which would involve multiplying two *N* × *N* matrices), our block-wise approach multiplied smaller sub-matrices (e.g. a *K* × *N* block with an *N* × *N* matrix, where *N* ≫ *K*), drastically reducing the total number of computations required. The size of each subnetwork block *K* was selected based on the memory and cores available per node to optimize performance without exceeding resource limits. We found that processing the data in blocks (saving approximately 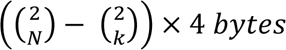 per block) reduced memory usage by hundreds of gigabytes per node and improved runtime by enabling parallel execution.

Another crucial optimization involved “neighbor contributions” for the block-wise GSN computation. In our parallel framework, each compute node processed a subset of SNPs (a block) but needed to account for connections to SNPs in other blocks. We addressed this by computing and storing the necessary cross-block contributions (essentially, the average adjacency contributions from SNPs outside the block, referring to the min(*s*_*i*_, *s*_*j*_) in Equation 1) once at the start of the computation. These neighbor contributions were then broadcast or made available to all nodes, so that each node could incorporate the external connectivity information into its local calculations without repeatedly recomputing or retrieving it from disk. By loading these contributions into memory once and reusing them, we avoided redundant I/O and computation, further reducing the runtime.

Other steps taken in *ESNA* to further reduce memory included developing a utility called *QuickMap* to efficiently index and retrieve values from the GSN without storing the full matrix in memory; we stored only the upper triangular half of the GSN matrix (or equivalently, the lower half) as a one-dimensional array. *QuickMap* contained logic to map any requested matrix index (*i*, *j*) to the position in this 1D array, allowing us to reconstruct the needed row or column on the fly. This method yielded approximately 50% memory savings, as only half of the matrix was stored. We parallelized the *QuickMap* retrieval computations across multiple CPU cores on a node to ensure that reconstructing rows/columns (when needed for the clustering step) was fast and did not become a bottleneck.

In essence, *QuickMap* traded a small amount of additional computation for a large reduction in memory usage. Specifically, given a *N* × *N* GSN matrix, *ESNA* required 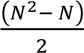 memory to store GSN information on a 1D array *A*. The function for reconstructing the *i*-th row of the original GSN matrix using *QuickMap* is shown as follows.

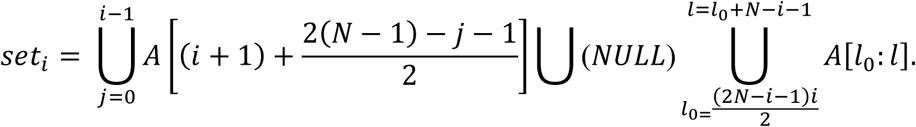

In the above equation, *set*_*i*_ represents the *i*-th row of the GSN, reconstructed from the 1D array *A*. This row was composed of three parts: 1) the elements from the *i*-th column in *A*, which had a length of *i*; 2) the diagonal element *Null*; and 3) the elements beyond the diagonal, with a length of *N* − *i* − 1. *QuickMap* achieves approximately 50% memory savings for non-sparse GSN matrices, as illustrated in Figure 1D.

## Results

### ESNA Computational Performance

We first executed *GLIDE* to calculate all pairwise SNP interaction statistics (t-scores for SNP-SNP interaction terms) using genome-wide data from the CHILD Study. This initial epistasis scan (775,613 SNPs) completed in 13 hours using 2 terabytes (TB) of RAM, producing the raw SNP-SNP similarity results. Next, we normalized the GLIDE output using min-max normalization and stored it as a sparse SNP similarity matrix in HDF5 format. This step took 11 hours and required up to 1 TB of memory. We then determined the optimal soft-thresholding power parameter (*λ* in Equation 1) for network construction using a custom adjacency function. This step was completed in around 1.5 hours on a CPU node with 650 GB of memory.

To construct the global SNP network (GSN) from a similarity matrix, we leveraged a distributed GPU computing strategy. The adjacency matrix was split into 1,292 submatrices of 600 rows each, with the remaining smaller tail block of 369 rows handled by a fifth node. The parallelization scheme, shown in Figure 1C, allowed us to construct the GSN in approximately 3 days using 5 GPU nodes (each with 175 GB RAM) simultaneously. Merging the submatrices into the full GSN using the QuickMap function took about 2 days and used less than 1TB of RAM, with GSN data stored on disk in float16 format. We then performed SNP clustering on the GSN in two stages. First, a customized K-means algorithm (inspired by the WGCNA *projectiveKMeans* function [45] and reimplemented in Python) was applied to pre-cluster the SNPs into preliminary groups, parallelized across 8 CPU cores (*mpi4py* [33] library, 1 TB memory). This K-means pre-clustering completed in 10 days. Second, we refined the preliminary clusters by applying hierarchical clustering with *DynamicTreeCut* (*linkage* and *cutreeHybrid*), which required 166 GB of memory and 1 hour of CPU time to produce the final modules.

### Pairwise SNP-SNP Epistatic Analysis

Genome-wide pairwise epistatic analysis using GLIDE initially assessed 775,613 annotated SNPs, identifying 1,792,446,238 SNP-SNP interactions that surpassed the significance threshold (t-score ≥ 3). Following quality control, including the removal of SNPs with missing (NaN) values, 775,569 SNPs were retained for further analysis. These interactions were assembled into a SNP similarity matrix and a corresponding denoised adjacency matrix, both with dimensions of 775,569 × 775,569. Despite the large scale of the dataset, the matrices remained highly sparse, with only approximately 0.3% of all possible SNP pairs exhibiting significant interaction (sparsity 99.7%), which was consistent with our expectation that only a small fraction of pairwise interactive SNPs would impact recurrent wheeze.

### Global SNP Network (GSN) Construction and Clustering of SNP Modules

We next converted the filtered pairwise similarity matrix into a higher-order network representation to capture extended epistatic relationships. Using the chosen power parameter *λ* = 7 from Equation 1, each pairwise similarity was transformed into an adjacency weight, constructing the Global SNP Network (GSN) comprising 775,569 nodes (SNPs). We then sought to identify groups of SNPs that formed highly connected network communities (putative epistatic modules). Using our two-stage clustering approach, we first applied parallel K-means to partition the SNPs into 128 preliminary clusters based on their GSN similarity profiles. We then performed hierarchical clustering within each preliminary cluster (using the DynamicTreeCut algorithm) to refine the community structure. This approach resulted in a final set of 914 distinct SNP modules spanning the 775,569 SNPs in the network.

### Module Associations and Annotations

To assess the phenotypic relevance of the identified SNP modules, we tested each module for association with the respiratory outcomes. We employed SKAT to evaluate the joint effect of variants in each module on the recurrent wheeze and childhood asthma phenotypes. To increase power, we incorporated prior knowledge by weighting SNPs according to their effect sizes (beta coefficients) from a large asthma GWAS [36]). Out of the 914 modules tested, 9 modules showed significant association with recurrent wheeze at a Bonferroni-adjusted significance threshold of 5.471×10^-5^ (0.05/914), summarized in **Table 1**. Collectively, they contained 72,700 SNPs, accounting for approximately 9.37% (72,700/775,569) of the total SNPs analyzed. Notably, 7 of these 9 wheeze-associated modules also displayed significant associations with asthma at age 5 (**Table 1**).

**Table 1.** SNP modules identified by ESNA and associated with recurrent wheeze and asthma.

| SNP Cluster | No. of SNPs | P (Recurrent wheeze) | P (Asthma) |
| --- | --- | --- | --- |
| **<u>Cluster686</u>** | 6406 | 1.309781E-08 | 2.485591E-05 |
| **<u>Cluster157</u>** | 4219 | 4.757537E-08 | 2.104057E-05 |
| **<u>Cluster416</u>** | 7574 | 6.355959E-08 | 9.831348E-07 |
| **<u>Cluster257</u>** | 12033 | 6.737406E-08 | 2.371874E-06 |
| **<u>Cluster504</u>** | 7407 | 9.310124E-08 | 4.164972E-06 |
| **<u>Cluster438</u>** | 3450 | 1.168849E-07 | 1.947806E-06 |
| Cluster290 | 5831 | 3.357418E-07 | 1.190439E-04 |
| **<u>Cluster95</u>** | 18764 | 1.954602E-05 | 1.532069E-05 |
| Cluster282 | 7016 | 3.452017E-05 | 1.340407E-02 |

Note: SNP modules associated with both asthma and recurrent wheeze are in bold and underlined, which surpassed the Bonferroni-adjusted significance threshold of $5.47 \times 10^{-5}$ (0.05/914). Nine modules were significantly associated with recurrent wheeze. Seven of these modules were also associated with asthma.

We performed extensive functional annotation of the SNP content and biological roles of these significant modules. Using the *Ensembl Variant Effect Predictor (VEP)* [37], SNPs in each module were mapped to their nearest genes and functional consequences. Gene-set enrichment analysis with *g:Profiler* [38] based on these gene symbols revealed significant enrichment in various biological pathways.

For the most prominent module, Cluster686, *g:Profiler* identified strong enrichment for Gene Ontology (GO) terms, particularly those related to nervous system development, cellular response to stimulus, and immune-related processes (Figure 2). Figure 2A presents an overview of all significantly enriched GO terms across biological process (BP), molecular function (MF), and cellular component (CC) categories, highlighting a subset of 43 highly enriched pathways passing a stringent threshold of −*log*_10_(*P*) = 16. Figure 2B focuses on the top 30 GO:BP pathways, shown in a dot-plot with size representing gene counts and color representing significance. Full *g:Profiler* results for Cluster686, including all enriched terms and statistics, are provided in **Supplemental Table 1.** Similar pathway enrichments were observed across other significant modules. Notably, Cluster686 contained a concentrated set of immune-related variants: 156 SNPs in this module were annotated with immune response functions in the *Gene Ontology* database [46]. Additionally, we found that 31 SNPs across the 9 significant modules overlapped with known childhood asthma loci reported in the *GWAS catalog* [47] (see **Supplemental Table 2**).

**Figure 2.**
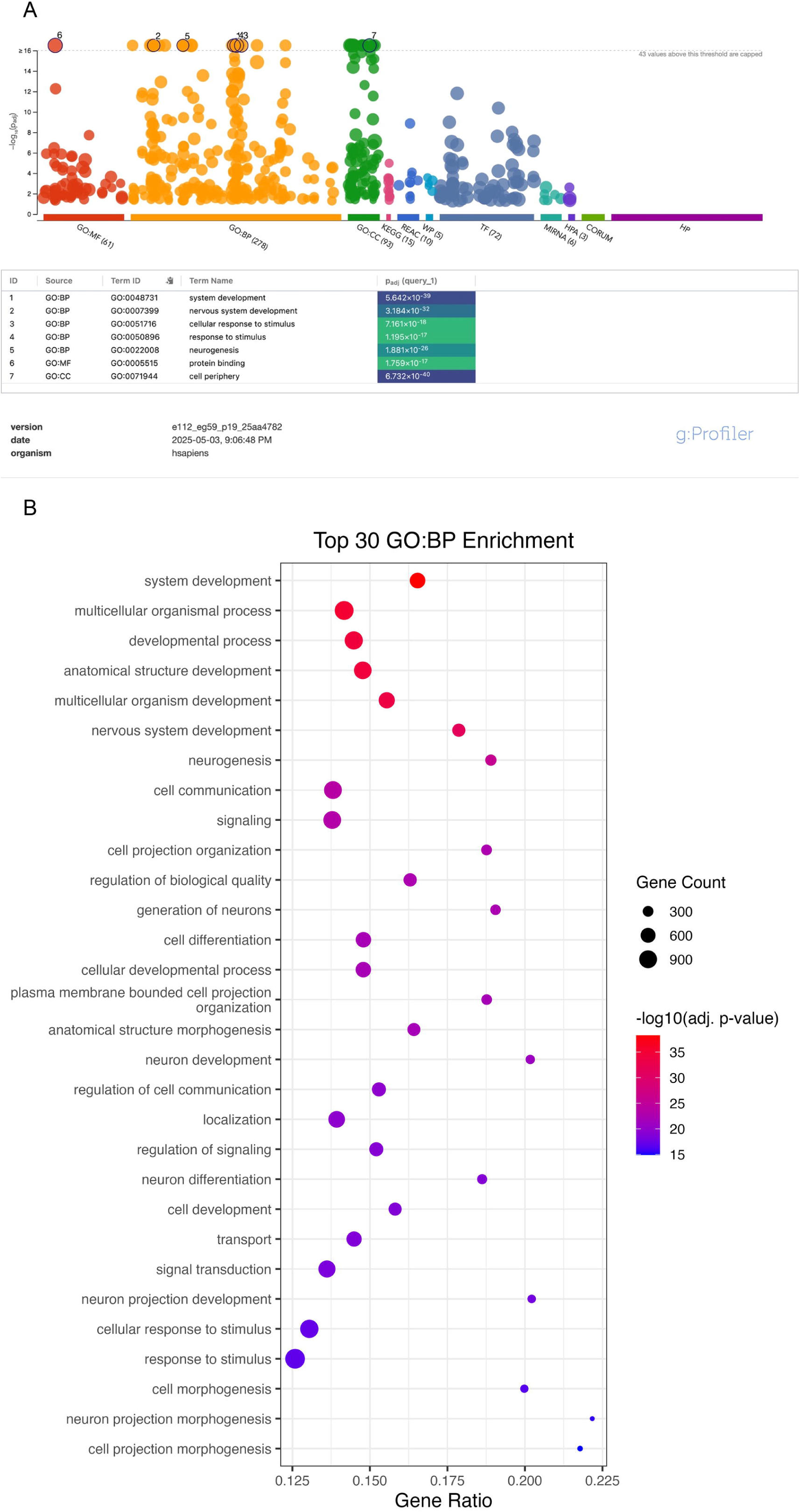
Functional gene enrichment analysis of the SNP network associated with recurrent wheeze and asthma highlights plausible biological processes. **(A)** Overview of all significantly enriched GO terms (GO:BP, GO:MF, and GO:CC) based on gene symbols input from Cluster686 using *g:Profiler*. The dashed horizontal line indicates a capped significance threshold at −*log*_10_(*pvalue*) = 16, highlighting highly enriched terms. A total of 43 GO terms passed this threshold, including the pathways in GO:BP, such as nervous system development (P=5.64×10*^-39^*), cellular response to stimulus (P=7.161×10*^-18^*), and response to stimulus (P=1.20×10^-17^). **(B)** Detailed dot plot of the top 30 GO:BP terms based on adjusted P. The x-axis represents the gene ratio (number of genes in the intersection divided by the total number of genes annotated to the term), the y-axis lists the pathway descriptions, dot size reflects the number of contributing genes, and dot color corresponds to statistical significance (−log₁₀ adjusted P).

Beyond functional enrichment, we examined the genomic distribution of SNPs within each associated module. The modules identified by ESNA tended to connect variants that spanned multiple genes or lay in non-coding regions. For example, Cluster686 comprised a large proportion of variants located in intergenic regions (e.g., 4,227 out of 6,406 SNPs), consistent with the fact that linkage disequilibrium pruning does not prioritize SNPs based on genomic location. Although this intergenic predominance was expected due to the nature of genome-wide variant distribution, we noted that Cluster95 contained a relatively higher proportion of intragenic SNPs compared to other modules. A full summary of the intergenic versus intragenic SNP proportions across the 9 modules is provided in **Supplemental Table 3**. These results are broadly consistent with the notion that epistatic networks often span regulatory or non-coding regions, supporting the hypothesis that non-additive genetic effects may arise from distributed elements beyond protein-coding genes.

### GWAS vs. ESNA

To compare our network-based findings with traditional single-variant analysis, we conducted a genome-wide association study (GWAS) of recurrent wheeze in the CHILD Study. GWAS identified 31 SNPs that reached genome-wide significance (P < 5×10^-8^) and 114 additional SNPs with suggestive significance (P < 1×10^-5^). All 31 of these top GWAS hits mapped to a single locus on chromosome 17. One of the top GWAS SNPs, rs2941519 (GWAS P = 3.97×10^-8^), was also found within the most significant ESNA module, Cluster686 (module P = 1.31×10⁻⁸).

To explore the broader concordance between ESNA modules and GWAS signals, we examined the overlap between the top 9 ESNA-derived SNP modules and the 145 suggestively associated SNPs from GWAS. There were 8 overlapping SNPs, including rs2941519, indicating limited direct concordance. The overlapping SNPs are listed in **Supplemental Table 4**.

Finally, to visualize the relationship between the network-derived findings and conventional GWAS results, we mapped the module SNPs onto the GWAS Manhattan plot. In Figure 3, the 72,700 SNPs belonging to the 9 significant modules are highlighted in red on the GWAS significance landscape. This visualization illustrates that the epistatic SNPs identified by ESNA are distributed across the genome, often in regions that do not contain genome-wide significant SNP signals.

**Figure 3.**
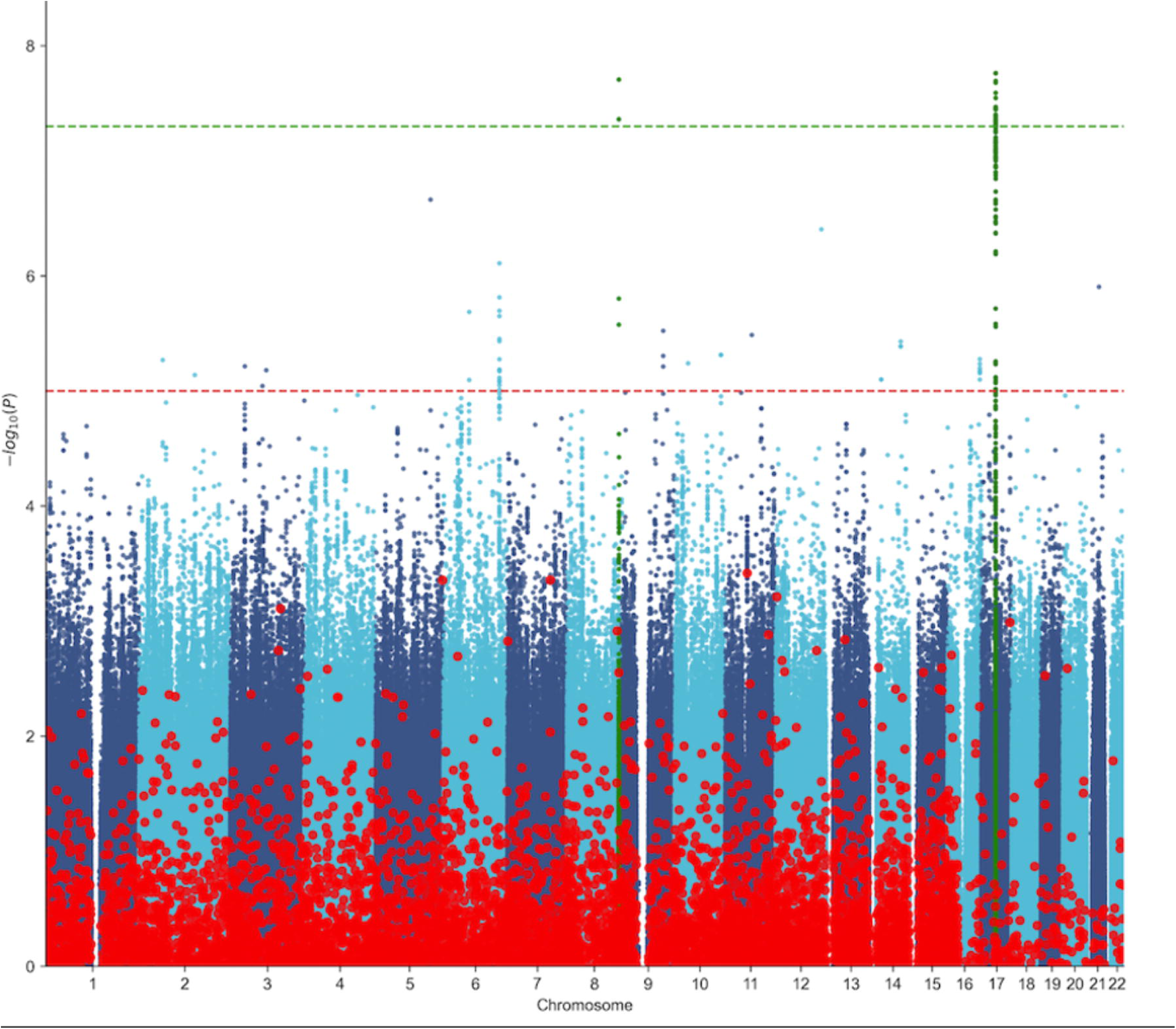
ESNA correlated modules captured additional SNPs with moderate associations with recurrent wheeze compared to GWAS results. The x-axis represents the chromosomal coordinates of SNPs; the y-axis shows statistical significance as −*log*_10_(*pvalue*). Horizontal lines indicate significance thresholds: red for suggestive (P = 1× 10⁻⁵) and green for genome-wide significance (P = 5×10⁻⁸). Red dots represent SNPs belonging to the top 9 significant ESNA module. Blue dots represent all 775,569 SNPs tested in both GWAS and ESNA.

## Discussion

In this study, we present ESNA (Epistatic SNP Network Analysis), a customized network-based computational framework designed to efficiently detect higher-order epistatic interactions from genome-wide SNP data. Whereas most existing epistasis detection methods are limited to identifying only pairwise SNP-SNP interactions, ESNA captures higher-order epistatic structures by constructing a global SNP network (GSN) and clustering interacting SNP modules. This is achieved through topological overlap-based network transformation of pairwise interaction scores that enables integration of indirect relationships across shared neighbors, thereby capturing higher-order interaction structure beyond isolated pairwise effects. Our methodological advancement addresses a major gap in genetic studies, where traditional approaches miss complex multi-locus interactions that likely contribute to the missing heritability of common traits and diseases.

Through the development of a homogeneous-computing framework and parallelized algorithm design, ESNA improves computational scalability, achieving significant reductions in runtime and memory usage. When applied to genomics data from a large Canadian birth cohort (CHILD study), ESNA successfully identified large clusters of interacting SNPs associated with recurrent wheeze and childhood asthma. Notably, while previous network-based tools such as *WISH* would require approximately two years to complete genome-wide analysis at this scale, *ESNA* completed the full analysis within 15 days, using optimized memory management strategies that reduced resource demands by over 50%. The ability to harness multiple GPU nodes through parallel computing further underscores ESNA’s practicality for real-world large-scale genomic studies. The ESNA pipeline is implemented as an end-to-end workflow, with publicly available code and documentation, to facilitate reproducibility and usability for large-scale genomic analyses.

Beyond computational improvements, *ESNA* addresses a gap in detecting interactive SNP effects often missed by conventional GWAS approaches. Traditional GWAS models evaluate the marginal effect of individual variants, leaving much of the heritability of complex traits unexplained. In contrast, *ESNA* uncovered SNP modules whose collective effects significantly contributed to phenotype variation, even when constituent SNPs lacked genome-wide significance. For example, rs2941519 was flagged both by single-SNP GWAS and as part of an ESNA-derived module (“Cluster686”), which was previously linked to ORMDL3 expression via eQTL analysis [48], and is one of the most replicated loci for childhood asthma [49]. Notably, *ESNA* also uncovered modules comprising SNPs with modest individual effects that, when considered together, revealed strong associations. These findings highlighted how *ESNA’s* results complement GWAS by uncovering higher-order genetic architecture underlying diseases like asthma.

Applying *ESNA* to the CHILD Cohort Study, we identified 9 SNP modules significantly associated with recurrent wheeze and asthma. Pathway enrichment revealed involvement in cellular stimulus and nervous system development, processes implicated in asthma pathophysiology. Prior studies highlighted how environmental triggers activated airway epithelial cells and afferent nerves, facilitating cell-cell communication in asthma [50]. Additionally, *Kistemaker et al.* [51] demonstrated the involvement of neural pathways, from the central nervous system to postganglionic neurons in airway walls, in regulating airway responses characteristic of asthma. *Ritz et al.* [52, 53] further emphasized the role of the central nervous system in modulating airway behavior in asthma, supporting the relevance of these pathways.

While *ESNA* provides a scalable and robust framework for genome-wide detection of higher-order epistasis, limitations remain. First, replications in external cohorts are necessary to validate the SNP modules associated with recurrent wheeze and asthma identified in this study. Second, although ESNA achieves substantial computational efficiency through parallel processing and memory optimization, further acceleration could be achieved by re-implementing key components in low-level programming languages such as C or Python’s *Cython* library [54]. Compiled execution and static typing offered by these languages would reduce interpretation overhead and optimize runtime performance. Third, while the two-stage clustering strategy (parallel K-means followed by *DynamicTreeCut*) effectively managed computational complexity for large datasets, it may be susceptible to local optima, a well-known challenge in unsupervised machine learning. Future enhancements could include dynamic monitoring to adaptively adjust clustering parameters or convergence criteria, mitigating the risk of suboptimal module detection. Addressing these limitations would further strengthen the robustness and generalizability of ESNA for broader applications in genetic epidemiology.

## Key Points

- Despite the success of genome-wide association studies (GWAS), efficient methods for detecting non-additive effects from higher-order variant interactions (epistasis) at a genome-wide scale remain limited.
- We introduce Epistatic SNP Network Analysis (ESNA), a network-based framework for detecting interactive SNP modules across the genome using a homogeneous parallel computing architecture.
- ESNA enables detection of higher-order SNP interactions beyond pairwise effects, uncovering polygenic signals that are often missed by traditional GWAS approaches.
- Compared to existing network-based methods, ESNA improves computational efficiency, reducing runtime by 48-fold and memory usage by approximately 50%.
- Application to 775,569 SNPs from the CHILD Cohort Study identified 9 modules associated with recurrent wheeze, 7 of which were also associated with asthma, with enrichment in biologically relevant pathways.

## Data Availability

Genotype and phenotype data from the CHILD Cohort Study are not publicly available due to participant privacy and ethical restrictions but may be made available upon reasonable request and with appropriate approvals from the CHILD Cohort Study and relevant institutional review boards.

## Acknowledgements

We are grateful to all the CHILD families who took part in this study, and the whole CHILD team, which includes interviewers, nurses, computer and laboratory technicians, clerical workers, research scientists, volunteers, managers, and receptionists. For a list of investigators and enrolling centres, visit www.childcohort.ca. The Canadian Institutes of Health Research (CIHR) and the Allergy, Genes and Environment Network of Centres of Excellence (AllerGen) provided core support for CHILD.

## References

1. Uffelmann, E., et al., Genome-wide association studies. Nature Reviews Methods Primers, 2021. 1(1): p. 59.

2. Visscher, P.M., et al., 10 Years of GWAS Discovery: Biology, Function, and Translation. Am J Hum Genet, 2017. 101(1): p. 5–22.

3. Wei, W.H., G. Hemani, and C.S. Haley, Detecting epistasis in human complex traits. Nat Rev Genet, 2014. 15(11): p. 722–33.

4. Niel, C., et al., A survey about methods dedicated to epistasis detection. Front Genet, 2015. 6: p. 285.

5. Wang, X., R.C. Elston, and X. Zhu, The meaning of interaction. Hum Hered, 2010. 70(4): p. 269–77.

6. van der Sijde, M.R., A. Ng, and J. Fu, Systems genetics: From GWAS to disease pathways. Biochim Biophys Acta, 2014. 1842(10): p. 1903–1909.

7. Ritchie, M.D., Finding the epistasis needles in the genome-wide haystack. Methods Mol Biol, 2015. 1253: p. 19–33.

8. Wan, X., et al., BOOST: A fast approach to detecting gene-gene interactions in genome-wide case-control studies. Am J Hum Genet, 2010. 87(3): p. 325–40.

9. Hemani, G., et al., EpiGPU: exhaustive pairwise epistasis scans parallelized on consumer level graphics cards. Bioinformatics, 2011. 27(11): p. 1462–5.

10. Liu, Y., et al., Genome-wide interaction-based association analysis identified multiple new susceptibility Loci for common diseases. PLoS Genet, 2011. 7(3): p. e1001338.

11. Gyenesei, A., et al., BiForce Toolbox: powerful high-throughput computational analysis of gene-gene interactions in genome-wide association studies. Nucleic Acids Res, 2012. 40(Web Server issue): p. W628-32.

12. Kam-Thong, T., et al., GLIDE: GPU-based linear regression for detection of epistasis. Hum Hered, 2012. 73(4): p. 220–36.

13. Carmelo, V.A.O., et al., WISH-R- a fast and efficient tool for construction of epistatic networks for complex traits and diseases. BMC Bioinformatics, 2018. 19(1): p. 277.

14. Saha, S., et al., Epi-MEIF: detecting higher order epistatic interactions for complex traits using mixed effect conditional inference forests. Nucleic Acids Res, 2022. 50(19): p. e114.

15. Graca, M., et al., Distributed transformer for high order epistasis detection in large-scale datasets. Sci Rep, 2024. 14(1): p. 14579.

16. Sun, L., et al., Epi-SSA: A novel epistasis detection method based on a multi-objective sparrow search algorithm. PLoS One, 2024. 19(10): p. e0311223.

17. Sun, Y., et al., MDSN: A Module Detection Method for Identifying Higher-order Epistatic Interactions. Genes (Basel), 2022. 13(12).

18. Subbarao, P., et al., The Canadian Healthy Infant Longitudinal Development (CHILD) Study: examining developmental origins of allergy and asthma. Thorax, 2015. 70(10): p. 998–1000.

19. Dharma, C., et al., Patterns of allergic sensitization and atopic dermatitis from 1 to 3 years: Effects on allergic diseases. Clin Exp Allergy, 2018. 48(1): p. 48–59.

20. Taussig, L.M., et al., Tucson Children’s Respiratory Study: 1980 to present. J Allergy Clin Immunol, 2003. 111(4): p. 661–75; quiz 676.

21. Dai, R., et al., Wheeze trajectories: Determinants and outcomes in the CHILD Cohort Study. J Allergy Clin Immunol, 2022. 149(6): p. 2153–2165.

22. Carr, T.F. and E. Bleecker, Asthma heterogeneity and severity. World Allergy Organ J, 2016. 9(1): p. 41.

23. Reyna, M.E., et al., Development of a Symptom-Based Tool for Screening of Children at High Risk of Preschool Asthma. JAMA Netw Open, 2022. 5(10): p. e2234714.

24. Comberiati, P. and C. Riggioni, Editorial comments on: "Persistence of asthma-like symptoms at early ages: A longitudinal twin study". Pediatr Allergy Immunol, 2022. 33(3): p. e13763.

25. Papi, A., et al., Asthma. Lancet, 2018. 391(10122): p. 783–800.

26. Moraes, T.J., et al., The Canadian healthy infant longitudinal development birth cohort study: biological samples and biobanking. Paediatr Perinat Epidemiol, 2015. 29(1): p. 84–92.

27. Ambalavanan, A., et al., Human milk oligosaccharides are associated with maternal genetics and respiratory health of human milk-fed children. Nat Commun, 2024. 15(1): p. 7735.

28. Purcell, S., et al., PLINK: a tool set for whole-genome association and population-based linkage analyses. Am J Hum Genet, 2007. 81(3): p. 559–75.

29. Laurie, C.C., et al., Quality control and quality assurance in genotypic data for genome-wide association studies. Genet Epidemiol, 2010. 34(6): p. 591–602.

30. Marees, A.T., et al., A tutorial on conducting genome-wide association studies: Quality control and statistical analysis. Int J Methods Psychiatr Res, 2018. 27(2): p. e1608.

31. Wu, M.C., et al., Rare-variant association testing for sequencing data with the sequence kernel association test. Am J Hum Genet, 2011. 89(1): p. 82–93.

32. Stuart, J.M., et al., A gene-coexpression network for global discovery of conserved genetic modules. Science, 2003. 302(5643): p. 249-55.

33. mpi4py, MPI for Python. https://mpi4py.readthedocs.io/en/stable/.

34. Langfelder, P. and S. Horvath, WGCNA: an R package for weighted correlation network analysis. BMC Bioinformatics, 2008. 9: p. 559.

35. Langfelder, P., B. Zhang, and S. Horvath, Defining clusters from a hierarchical cluster tree: the Dynamic Tree Cut package for R. Bioinformatics, 2008. 24(5): p. 719–20.

36. Pividori, M., et al., Shared and distinct genetic risk factors for childhood-onset and adult-onset asthma: genome-wide and transcriptome-wide studies. Lancet Respir Med, 2019. 7(6): p. 509–522.

37. McLaren, W., et al., The Ensembl Variant Effect Predictor. Genome Biol, 2016. 17(1): p. 122.

38. Raudvere, U., et al., g:Profiler: a web server for functional enrichment analysis and conversions of gene lists (2019 update). Nucleic Acids Res, 2019. 47(W1): p. W191–W198.

39. CACC, The Centre for Advanced Computing Canada. https://cac.queensu.ca.

40. Mason, C.E., et al., Standardizing the next generation of bioinformatics software development with BioHDF (HDF5). Adv Exp Med Biol, 2010. 680: p. 693–700.

41. scipy.sparse, Sparse matrices operations in Python. https://docs.scipy.org/doc/scipy/reference/sparse.html.

42. Golub, G.H.C.F.V.L., Matrix Computations (3rd ed.). 1996.

43. CuPy, NumPy/SciPy-compatible Array Library for GPU-accelerated Computing with Python. https://cupy.dev.

44. S., C., CUDA Programming: A Developer’s Guide to Parallel Computing with GPUs. USA: Morgan Kaufmann Publishers Inc., 2012. 1st ed.

45. function, p., Blockwise module analysis_wgcna_R. https://github.com/cran/WGCNA/blob/master/R/blockwiseModulesC.R.

46. Ashburner, M., et al., Gene ontology: tool for the unification of biology. The Gene Ontology Consortium. Nat Genet, 2000. 25(1): p. 25–9.

47. Sollis, E., et al., The NHGRI-EBI GWAS Catalog: knowledgebase and deposition resource. Nucleic Acids Res, 2023. 51(D1): p. D977–D985.

48. Li, X., et al., Genetic analyses identify GSDMB associated with asthma severity, exacerbations, and antiviral pathways. J Allergy Clin Immunol, 2021. 147(3): p. 894–909.

49. Moffatt, M.F., et al., Genetic variants regulating ORMDL3 expression contribute to the risk of childhood asthma. Nature, 2007. 448(7152): p. 470-3.

50. Erle, D.J. and D. Sheppard, The cell biology of asthma. J Cell Biol, 2014. 205(5): p. 621–31.

51. Kistemaker, L.E.M. and Y.S. Prakash, Airway Innervation and Plasticity in Asthma. Physiology (Bethesda), 2019. 34(4): p. 283–298.

52. Ritz, T., et al., Central nervous system signatures of affect in asthma: associations with emotion-induced bronchoconstriction, airway inflammation, and asthma control. J Appl Physiol (1985), 2019. 126(6): p. 1725-1736.

53. Ritz, T., et al., Airway response to emotional stimuli in asthma: the role of the cholinergic pathway. J Appl Physiol (1985), 2010. 108(6): p. 1542-9.

54. Cython, An optimising static compiler for both the Python programming language and the extended Cython programming language. https://cython.org.

